# Clinical, Echocardiographic, and Longitudinal Characteristics Associated with Heart Failure with Improved Ejection Fraction

**DOI:** 10.1101/2023.08.25.23294644

**Authors:** Erick Romero, Alexander Francois Baltodano, Paulo Rocha, Camryn Sellers-Porter, Dev Jaydeep Patel, Saad Soroya, Julie Bidwell, Imo Ebong, Michael Gibson, David A. Liem, Shirin Jimenez, Heejung Bang, Padmini Sirish, Nipavan Chiamvimonvat, Javier E. Lopez, Martin Cadeiras

**Affiliations:** Division of Cardiovascular Medicine, UC Davis Medical Center, Sacramento, CA; University of California Davis, School of Medicine, Sacramento, CA; Betty Irene Moore School of Nursing, University of California Davis, Sacramento, CA; Division of Biostatistics, Department of Public Health Sciences, University of California, Davis, Davis, CA; Division of Cardiovascular Medicine, Department of Internal Medicine, University of California, Davis, Davis, CA

**Author notes:** **Address for Correspondence**: Martin Cadeiras and Erick Romero, Division of Cardiovascular Medicine, UC Davis Medical Center, 4301 X St. Sacramento, 95817 CA, USA. **Disclosures:** The authors declare no conflicts of interest. Funding sources were not involved in the collection and interpretation of data, preparation of the manuscript, and decisions for publication. All authors listed have made a substantial, direct, and intellectual contribution to the work and approved it for publication. This study was reviewed and approved by UC Davis IRB. All data used were de-identified prior to analysis.

**Keywords:** HFrEF, HFimpEF, echocardiogram, trajectories, longitudinal

## Abstract

**Background:** Heart failure (HF) with improved ejection fraction (HFimpEF) has better outcomes than HF with reduced ejection fraction (HFrEF). However, factors contributing to HFimpEF remain unclear. This study aimed to evaluate clinical and longitudinal characteristics associated with subsequent HFimpEF.

**Methods:** This was a single-center retrospective HFrEF cohort study. Data were collected from 2014 to 2022. Patients with HFrEF were identified using ICD codes, echocardiographic data, and natriuretic peptide levels. The main endpoints were HFimpEF (defined as ejection fraction >40% at ≥3 months with ≥10% increase) and mortality. Cox proportional hazards and mixed effects models were used for analyses.

**Results:** The study included 1307 HFrEF patients with a median follow-up of 16.3 months (IQR 8.0-30.6). The median age was 65 years; 68% were male while 57% were white. On follow-up, 39% (n=506) developed HFimpEF, while 61% (n=801) had persistent HFrEF. A multivariate Cox regression model identified sex, race comorbidities, echocardiographic, and natriuretic peptide as significant covariates of HFimpEF (*p*<0.05). The HFimpEF group had better survival compared to the persistent HFrEF group (*p*<0.001). Echocardiographic and laboratory trajectories differed between groups.

**Conclusion:** In this HFrEF cohort, 39% transitioned to HFimpEF and approximately 50% met the definition within the first 12 months. In a HFimpEF model, sex, comorbidities, echocardiographic parameters, and natriuretic peptide were associated with subsequent HFimpEF. The model has the potential to identify patients at risk of subsequent persistent or improved HFrEF, thus informing the design and implementation of targeted quality-of-care improvement interventions.

## INTRODUCTION

For a subset of individuals with heart failure (HF) with reduced ejection fraction (HFrEF), some improvement in ejection fraction occurs. In order to be formally classified as heart failure with improved ejection fraction (HFimpEF), however, the individual must have a baseline left ventricular ejection fraction (LVEF) of ≤40% and subsequent LVEF >40% at ≥3 months, with ≥10% absolute increase.^1–3^

The prognosis and outcomes for HFimpEF tend to be better as compared to persistent HFrEF. Persons with HFimpEF have significantly lower rates of mortality, cardiac hospitalizations, all-cause hospitalizations, and composite events.^4–9^ Moreover, HFimpEF patients have lower rates of cardiac transplantation, left ventricular assist device implantations, and a significant enhancement in health-related quality of life.^10^ However, the underlying mechanisms and factors leading to HFimpEF are not yet fully understood. The improvement in ejection fraction seen in HFimpEF can be due to the use of evidence-based medical therapy, device therapies, spontaneous improvement, or a combination of them.^11,12,1^ Nonetheless, specific patient clinical characteristics or treatments that contribute to HFimpEF are not always clear.

Factors such as the etiology of the initial injury, female sex, and non-ischemic comorbidities have been related to ejection fraction improvement and subsequent clinical outcomes.^7^ Echocardiographic data on HFimpEF patients have highlighted an initial better left ventricular dimensions, and diastolic function compared to persistent HFrEF patients.^8,9^ However, the absence of a widespread consensus on the clinical, laboratory, echocardiographic, and therapy factors associated with ejection fraction improvement remains a challenge. A better understanding of those factors could help clinicians to predict which individuals are at risk of persistent HFrEF or HFimpEF. This can inform and design management strategies for this complex population. Therefore, the objective of this study is to evaluate clinical and longitudinal factors and the associated with subsequent HFimpEF development.

## METHODS

### Study design and cohort data source

This was a single center retrospective cohort study conducted at the University of California, Davis Medical Center. The study was approved by the university’s Institutional Review Board, and all data used were de-identified prior to analysis.

Data was collected through the institution’s electronic health record data warehouse between January 2014 and December 2022. The study population consisted of adult patients (≥18 years) diagnosed with HFrEF. The HFrEF cohort inclusion criteria included ICD HF codes (ICD-9 or ICD-10), LVEF ≤40%, and Brain Natriuretic Peptide (BNP) ≥100 pg/mL. Diagnostic accuracy was evaluated on a random sub-sample (n=200) of the cohort and compared to physician chart review. Excluded were patients who had ICD codes for cardiac transplant or left ventricular assist device implantation, those who had only one LVEF value in the electronic medical record, and those with less than a three-month interval between their first and second evaluation of LVEF.

The demographic information extracted consisted of self-reported sex, race, and ethnicity. Baseline laboratory, echocardiogram, and electrocardiogram characteristics were defined as the first value within 90 days after the first LVEF ≤40% or the first available. Comorbidities were defined as diagnoses prior to the first LVEF ≤40% date, and validated comorbidity identification methodologies were used.^13,14^ For this study, we defined guideline directed medical therapy utilization as the prescription of medications within 6 months prior and within 3 months after the first LVEF ≤40% date. Data of medication of reconciliation was used to represent most complete possible information. The medications included renin-angiotensin-system inhibitor (RASi), β-blockers, mineralocorticoid receptor antagonist and sodium glucose co-transporter 2 inhibitors. The RASi category comprised angiotensin-converting enzyme inhibitor, angiotensin receptor blocker, and angiotensin receptor neprilysin inhibitor. Patients were placed into one of three dosing categories based on their recorded prescription: either none, <50%, or ≥50% of guideline directed therapy target dosing.^15^

Longitudinal laboratory, electrocardiogram, and echocardiogram variables were extracted, starting from the first LVEF ≤40% until the last available data point on follow-up. Data on all-cause mortality was obtained from the clinical data warehouse, with the latest data available within the study timeline being extracted.

### Heart failure cohort definitions

The cohort was stratified into two groups: one group consisting of persons with HF who had an improvement in ejection fraction (HFimpEF) and a second group consisting of persons who did not experience an improvement, but maintained persistently reduced ejection fraction (persistent HFrEF). We applied the following criterion to define HFimpEF: an initial LVEF of ≤40%, followed by a subsequent LVEF measurement of >40% at least 3 months later, with a minimum absolute improvement of 10%.^1–3^ Those who did not meet these criteria were categorized as persistent HFrEF patients.

### Endpoints

Endpoints were analyzed in a time-to-event fashion. The primary endpoint was the development of HFimpEF, and all-cause mortality was the secondary endpoint. For patients who did not experience HFimpEF, the last LVEF captured was considered the last follow up (so censoring). For the mortality outcome analysis, the last LVEF was considered the last follow up.

### Statistical analyses

Descriptive statistics were used to summarize baseline characteristics. Between-group comparisons were performed using Student’s t-test or Wilcoxon signed-rank test for continuous variables, and chi-squared or Fisher exact test for categorial variables as appropriate. In a complete-case analysis fashion, we first conducted univariate Cox regression analyses for each variable for the HFimpEF primary endpoint. Variables with *p<*0.20 in univariate analyses were included in the multivariate Cox regression model. The final multivariate model was selected using backward elimination with a retention threshold of *p*<0.05. For the multivariate model, collinearity between variables was tested using correlation analyses, with a correlation coefficient threshold of *r =* ±0.50. The robustness of the results and missing data were evaluated by performing a sensitivity analysis with multiple imputation (*m*=50), followed by pooling the derived parameter estimates and associated standard errors. Kaplan-Meier survival curves and the log-rank test were used to compare overall survival (secondary endpoint) between the HFimpEF and persistent HFrEF groups. The trajectories of echocardiogram and laboratory parameters over time were plotted using penalized B-spline curves. Linear mixed models for longitudinal data were performed to assess changes between groups over time. Analyses were performed using SAS software version 9.4 (SAS institute, Cary, NC, USA).

## RESULTS

### Study cohort population

The cohort identification criteria had a specificity of 0.96 and a sensitivity of 0.60 as compared with physician chart review (**Supplemental Table S1**). Patients were followed for a median of 16.3 months (IQR 8.0-30.6). Over the course of follow up, a total of n=506 (39%) patients developed HFimpEF (50% within the first year), while n=801 (61%) had persistent HFrEF (**Figure 1 and Supplemental Figure S1**).

**Figure 1.**
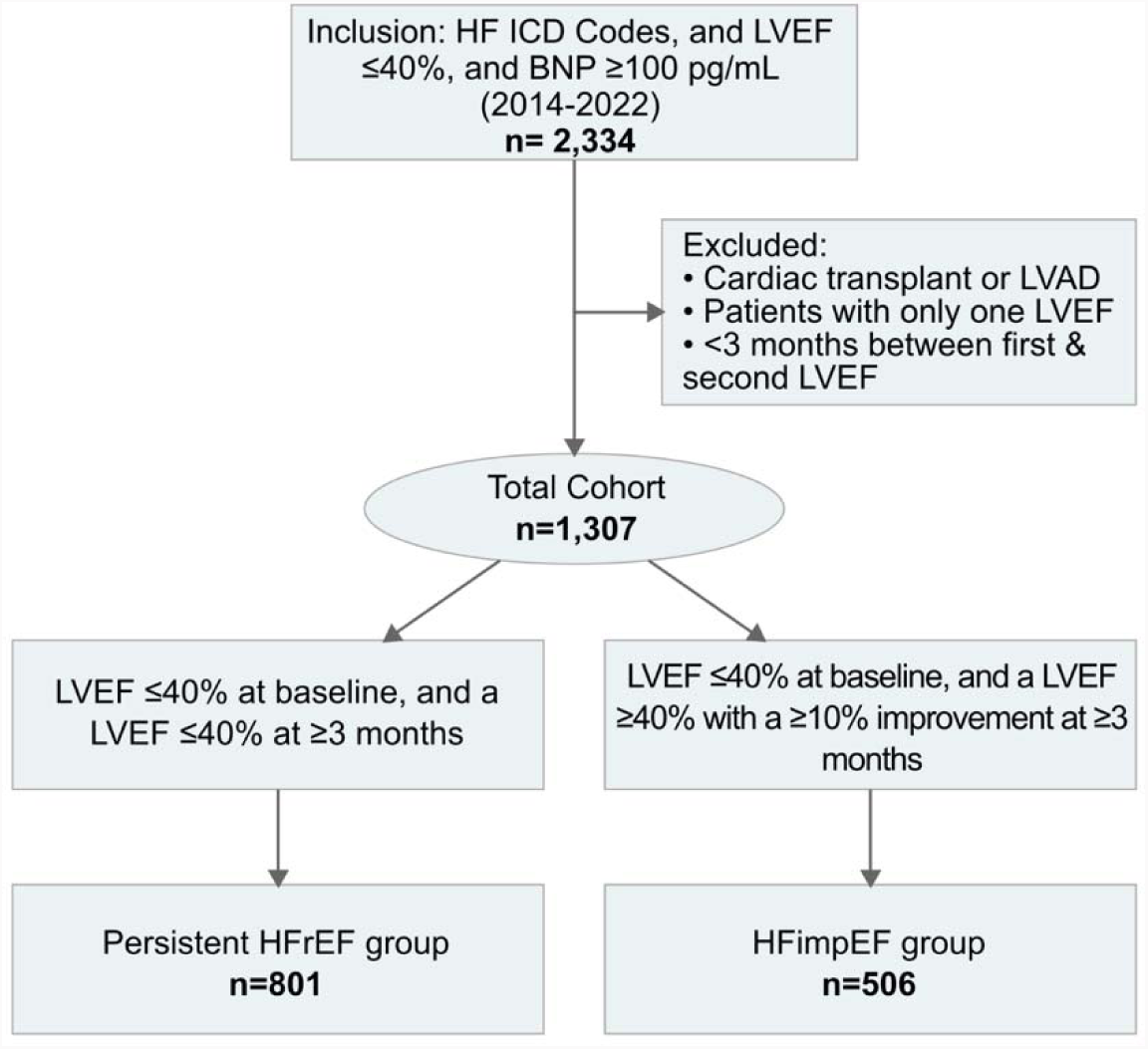
Cohort identification workflow.

Between-group comparisons of patient baseline characteristics are displayed in **Table 1**. The HFimpEF group consisted of older patients with a higher percentage of females as compared to the persistent HFrEF group. Additionally, the HFimpEF group exhibited a higher heart rate, body mass index, higher prevalence of hypertension and atrial fibrillation, while displaying a lower prevalence of coronary artery disease. The HFimpEF group also had lower BNP and left ventricular internal dimension values, along with higher LVEF, posterior wall, and interventricular septum thickness. The guideline directed therapy utilization of RASi and β-blockers at ≥50% target doses were more frequent in the HFimpEF group. However, the persistent HFrEF group displayed an overall higher frequency in the use of RASi and β-blockers.

**Table 1.** Patient baseline characteristics at the time of first LVEF ≤40%.

| Characteristics | Median (IQR) or No. (%) |  |  | P-Value<sup>a</sup> | n |
| --- | --- | --- | --- | --- | --- |
|  | All (N=1307) | Persistent HFrEF (N=801) | HFimpEF (N=506) |  |  |
| Age, years | 65 (55-75) | 63 (54-74) | 66 (58-76) | <.001 | 1307 |
| **Sex** |  |  |  | <.001 | 1307 |
| Male | 887 (67.9) | 582 (72.7) | 305 (60.3) |  |  |
| **Race** |  |  |  | 0.157 | 1307 |
| White | 742 (56.8) | 439 (54.8) | 303 (59.9) |  |  |
| Black | 229 (17.5) | 155 (19.4) | 74 (14.6) |  |  |
| Asian | 86 (6.6) | 49 (6.1) | 37 (7.3) |  |  |
| Native American/Hawaiian | 41 (3.1) | 25 (3.1) | 16 (3.2) |  |  |
| Other | 200 (15.3) | 129 (16.1) | 71 (14.0) |  |  |
| Unavailable | 9 (0.7) | 4 (0.5) | 5 (1.0) |  |  |
| **Ethnicity** |  |  |  | 0.910 | 1307 |
| Hispanic | 173 (13.2) | 108 (13.5) | 65 (12.9) |  |  |
| Non-Hispanic | 1128 (86.3) | 689 (86.0) | 439 (86.8) |  |  |
| Unavailable | 6 (0.5) | 4 (0.5) | 2 (0.4) |  |  |
| Heart rate, b.p.m. | 88 (75-102) | 86 (73-100) | 90 (77-105) | <.001 | 1306 |
| **Blood pressure, mm Hg** |  |  |  |  |  |
| Systolic | 127 (112-143) | 127 (112-142) | 127 (112-145) | 0.885 | 1307 |
| Diastolic | 77 (67-90) | 76 (67-90) | 78 (67-90) | 0.345 | 1307 |
| MAP | 94.3 (82.7-106.7) | 93.7 (82.7-106) | 95.3 (83.3-107) | 0.395 | 1307 |
| Weight, Kg | 83.5 (71.2-101.1) | 83.1 (70.6-98.7) | 84.1 (72.4-103.5) | 0.104 | 1306 |
| Body mass index | 28.2 (24.5-32.9) | 27.9 (24.2-32.2) | 28.7 (24.9-33.7) | **0.024** | 1172 |
| **Medical History** |  |  |  |  | 1295 |
| Hypertension | 967 (74.7) | 575 (72.4) | 392 (78.2) | **0.019** |  |
| Diabetes | 568 (43.9) | 347 (43.7) | 221 (44.1) | 0.885 |  |
| Hyperlipidemia | 574 (44.3) | 330 (41.6) | 244 (48.7) | **0.012** |  |
| Coronary artery disease | 478 (36.9) | 318 (40.1) | 160 (31.9) | **0.003** |  |
| Atrial fibrillation | 436 (33.7) | 231 (29.1) | 205 (40.9) | <.001 |  |
| Chronic kidney disease | 340 (26.3) | 210 (26.5) | 130 (25.9) | 0.842 |  |
| **Laboratory** |  |  |  |  |  |
| BNP, pg/mL | 623 (265-1330) | 750 (323-1521) | 451.5 (209-926) | <.001 | 1307 |
| NT-proBNP, pg/mL | 1110 (326-2914) | 1242 (431-3339) | 874 (218-2852) | 0.214 | 148 |
| Sodium, mEq/L | 138 (135-139) | 138 (135-139) | 138 (136-140) | 0.067 | 1306 |
| Potassium, mEq/L | 4 (3.7-4.4) | 4 (3.7-4.4) | 4 (3.7-4.4) | 0.813 | 1306 |
| Creatinine, mg/dL | 1.2 (0.9-1.5) | 1.2 (0.9-1.5) | 1.2 (0.9-1.5) | 0.952 | 1306 |
| eGFR, mL/min/1.73 m<sup>2</sup> | 56 (46-60) | 57 (47-61) | 55 (45-60) | **0.048** | 1232 |
| **Echocardiogram** |  |  |  |  |  |
| LVEF % | 30 (25-40) | 30 (20-35) | 35 (30-40) | <.001 | 1307 |
| IVSd | 1.2 (1-1.3) | 1.2 (1-1.3) | 1.2 (1.1-1.4) | <.001 | 1307 |
| LVIDd | 5.6 (5.1-6.2) | 5.8 (5.3-6.4) | 5.4 (4.9-5.8) | <.001 | 1306 |
| LVIDs | 4.7 (4.1-5.4) | 4.9 (4.3-5.6) | 4.4 (3.8-4.9) | <.001 | 1305 |
| PASP | 41 (30.5-50) | 41.5 (31.2-50.4) | 40 (29.7-49.7) | 0.293 | 1240 |
| PW | 1.2 (1-1.3) | 1.1 (1-1.3) | 1.2 (1-1.3) | <.001 | 1307 |
| TAPSE | 1.8 (1.4-2.1) | 1.8 (1.4-2.1) | 1.8 (1.4-2.1) | 0.818 | 1283 |
| **Electrocardiogram** |  |  |  |  |  |
| QTc, ms | 498 (472-528) | 499 (474-528) | 495.5 (470-527) | 0.161 | 1287 |
| **GDMT & target dose<sup>b</sup>** |  |  |  |  | 1059 |
| RASi<sup>c</sup> |  |  |  | **0.004** |  |
| none | 188 (17.8) | 109 (16.9) | 79 (19.1) |  |  |
| <50% target | 435 (41.1) | 291 (45.1) | 144 (34.9) |  |  |
| ≥50% target | 436 (41.2) | 246 (38.1) | 190 (46) |  |  |
| β-Blocker |  |  |  | **<.001** |  |
| none | 406 (38.3) | 214 (33.1) | 192 (46.5) |  |  |
| <50% target | 349 (33) | 255 (39.5) | 94 (22.8) |  |  |
| ≥50% target | 304 (28.7) | 177 (27.4) | 127 (30.8) |  |  |
| MRA |  |  |  | 0.654 |  |
| none | 694 (65.5) | 430 (66.6) | 264 (63.9) |  |  |
| <50% target | 2 (0.2) | 1 (0.2) | 1 (0.2) |  |  |
| ≥50% target | 363 (34.3) | 215 (33.3) | 148 (35.8) |  |  |
| SGLT2i | 70 (6.6) | 48 (7.4) | 22 (5.3) | 0.179 |  |
| GDMT Triple therapy | 178 (16.8) | 120 (18.6) | 58 (14) | 0.054 |  |
| GDMT Triple, ≥50% target | 60 (5.7) | 34 (5.3) | 26 (6.3) | 0.479 |  |

<sup>a</sup>P-Values comparing HFimPEF vs. persistent HFrEF.
<sup>b</sup>Target doses of guideline directed medical therapy (GDMT) as recommended in treatment guidelines.
<sup>c</sup>RASi consisted of angiotensin-converting enzyme inhibitor, angiotensin receptor blocker, angiotensin receptor neprilysin inhibitor.
**Abbreviations:** BNP= B-type natriuretic peptide; eGFR= estimated glomerular filtration rate; GDMT= guideline-directed medical therapy; IVSd= Interventricular septum thickness at end-diastole; LVEDd= left ventricle end diastolic diameter; LVEF= left ventricular ejection fraction; LVIDd= left ventricular internal dimension at end -diastole; LVIDs= left ventricular internal dimension at end -systole; MAP= mean arterial pressure; MRA= mineralocorticoid receptor antagonist; NT-proBNP= N-terminal (NT)-pro hormone BNP; PASP= pulmonary artery systolic pressure; PW= left ventricular posterior wall; QTc= QT corrected for heart rate; RASi= renin-angiotensin-system inhibitor; SGLT2i= Sodium glucose co-transporter 2 inhibitors; TAPSE= tricuspid annular plane systolic excursion.

### The HFimpEF primary endpoint and the secondary mortality endpoint

Results derived from the univariate analyses and subsequent multivariate Cox regression model for the primary HFimpEF endpoint are shown in **Table 2**. In the multivariate model, significant (*p*<0.05) baseline characteristics associated with HFimpEF included female sex, atrial fibrillation, elevated heart rate, higher first LVEF, and increased thickness of the interventricular septum thickness at end-diastole (IVSd). Covariates associated with persistent HFrEF included Black race, coronary artery disease, higher levels of BNP, increased left ventricular internal dimension (LVID) at end-diastole, and using β-blockers at <50% of the target dose (results summarized in **Figure 2**). The HFimpEF multivariate model had an acceptable predicting accuracy with a C-statistic of 0.68. The sensitivity analyses produced similar results (i.e., all predictors retained statistical significance). However, Black race did not reach significance levels (*p*>0.05), suggesting a weak association with the outcome.

**Figure 2.**
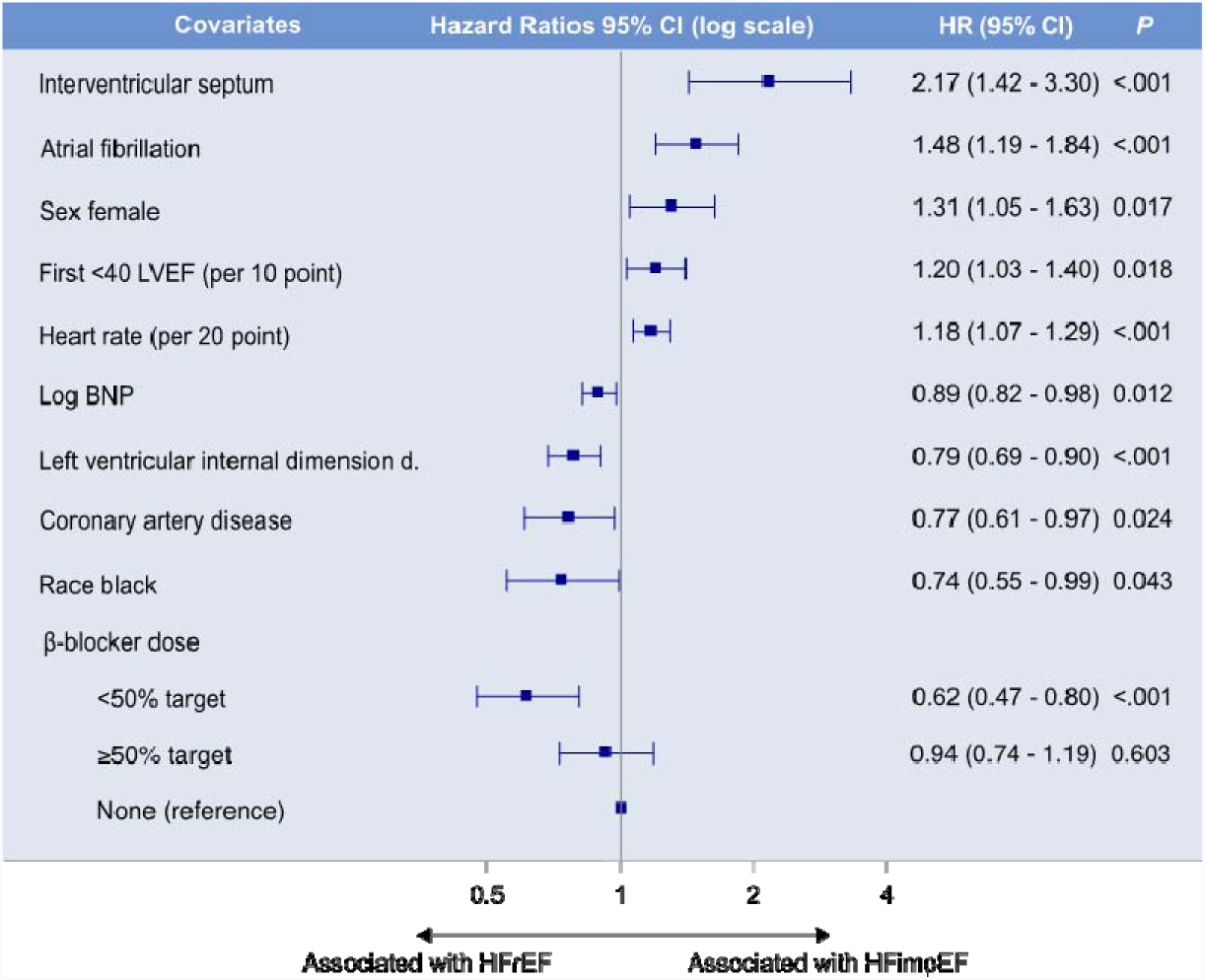
Summary of the multivariate Cox regression model for the HFimpEF primary outcome. A backward-elimination algorithm was utilized to select the final multivariable model, with a retention threshold of *p*≤.05. See Table 2 for details.

**Table 2.** Cox regression analyses for the of HFimpEF, primary endpoint.

| Variable | Univariate models |  | Multivariate model<sup>a</sup> |  |
| --- | --- | --- | --- | --- |
|  | HR (95% CI) | P value | HR (95% CI) | P value |
| Age at start, years | 1.01 (1.01-1.02) | <.001 | . | . |
| Female | 1.54 (1.29-1.84) | <.001 | 1.31 (1.05-1.63) | **0.017** |
| Race | . | . | . | . |
| Asian | 1.12 (0.80-1.58) | 0.504 | . | . |
| Black<sup>b</sup> | 0.76 (0.59-0.98) | 0.032 | 0.74 (0.55-0.99) | **0.043** |
| Native American/Hawaiian | 1.18 (0.71-1.94) | 0.531 | . | . |
| Other | 0.85 (0.66-1.10) | 0.227 | . | . |
| Unavailable | 1.16 (0.48-2.80) | 0.746 | . | . |
| White | Reference | . | . | . |
| Atrial fibrillation | 1.54 (1.29-1.84) | <.001 | 1.48 (1.19-1.84) | **<.001** |
| Hypertension | 1.28 (1.03-1.58) | 0.024 | . | . |
| Coronary artery disease | 0.81 (0.67-0.98) | 0.028 | 0.77 (0.61-0.97) | **0.024** |
| Hyperlipidemia<sup>c</sup> | 1.24 (1.04-1.48) | 0.016 | . | . |
| Heart rate (per 20-point increase) | 1.15 (1.06-1.25) | <.001 | 1.18 (1.07-1.29) | **<.001** |
| β-blocker | . | . | . | . |
| ≥50% | 0.82 (0.66-1.02) | 0.075 | 0.94 (0.74-1.19) | 0.603 |
| <50% | 0.49 (0.38-0.63) | <.001 | 0.62 (0.47-0.80) | **<.001** |
| None | Reference | . | . | . |
| Log BNP<sup>d</sup> | 0.87 (0.81-0.93) | <.001 | 0.89 (0.82-0.98) | **0.012** |
| GFR | 0.99 (0.98-1.00) | 0.005 | . | . |
| First LVEF ≤40 (per 10-point increase) | 1.46 (1.30-1.64) | <.001 | 1.20 (1.03-1.40) | **0.018** |
| LVIDd<sup>c</sup> | 0.65 (0.58-0.72) | <.001 | 0.79 (0.69-0.90) | **<.001** |
| IVSd<sup>c</sup> | 2.12 (1.52-2.94) | <.001 | 2.17 (1.42-3.30) | **<.001** |

<sup>a</sup>Predictors from the univariate analysis that showed a p-value of ≤.20 were subsequently incorporated into a multivariate analysis. A backward-elimination algorithm was utilized to select the final multivariable model, with a retention threshold of p≤05. The multivariate model had a C-statistic of 0.68.
<sup>b</sup>In the multivariate analysis, race was categorized as black or other.
<sup>c</sup>Hyperlipidemia was omitted in the multivariate model due to possible overlapping effect with coronary artery disease. PW and LVIDs were excluded due to the correlation coefficient >0.5 with IVSd and LVIDd respectively.
<sup>d</sup>The variable has been log-transformed.
**Abbreviations:** BNP= B-type natriuretic peptide; eGFR= estimated glomerular filtration rate; GDMT= guideline-directed medical therapy; IVSd= Interventricular septum thickness at end-diastole; LVEDd= left ventricle end diastolic diameter; LVEF= left ventricular ejection fraction; LVIDd= left ventricular internal dimension at end -diastole; LVIDs= left ventricular internal dimension at end -systole; MAP= mean arterial pressure; MRA= mineralocorticoid receptor antagonist; NT-proBNP= N-terminal (NT)-pro hormone BNP; PASP= pulmonary artery systolic pressure; PW= left ventricular posterior wall; QTc= QT corrected for heart rate; RASi= renin-angiotensin-system inhibitor; TAPSE= tricuspid Annular Plane Systolic Excursion.

For the secondary endpoint of all-cause mortality, the HFimpEF group had a median survival of 80 months (95% confidence interval: 74.4–93.7), while the persistent HFrEF group had a median survival of 62 months (95% confidence interval: 55.6–74.2). **Figure 3** depicts the Kaplan-Meier survival curves, indicating a significantly higher survival in the HFimpEF group (log-rank *p*<0.001).

**Figure 3.**
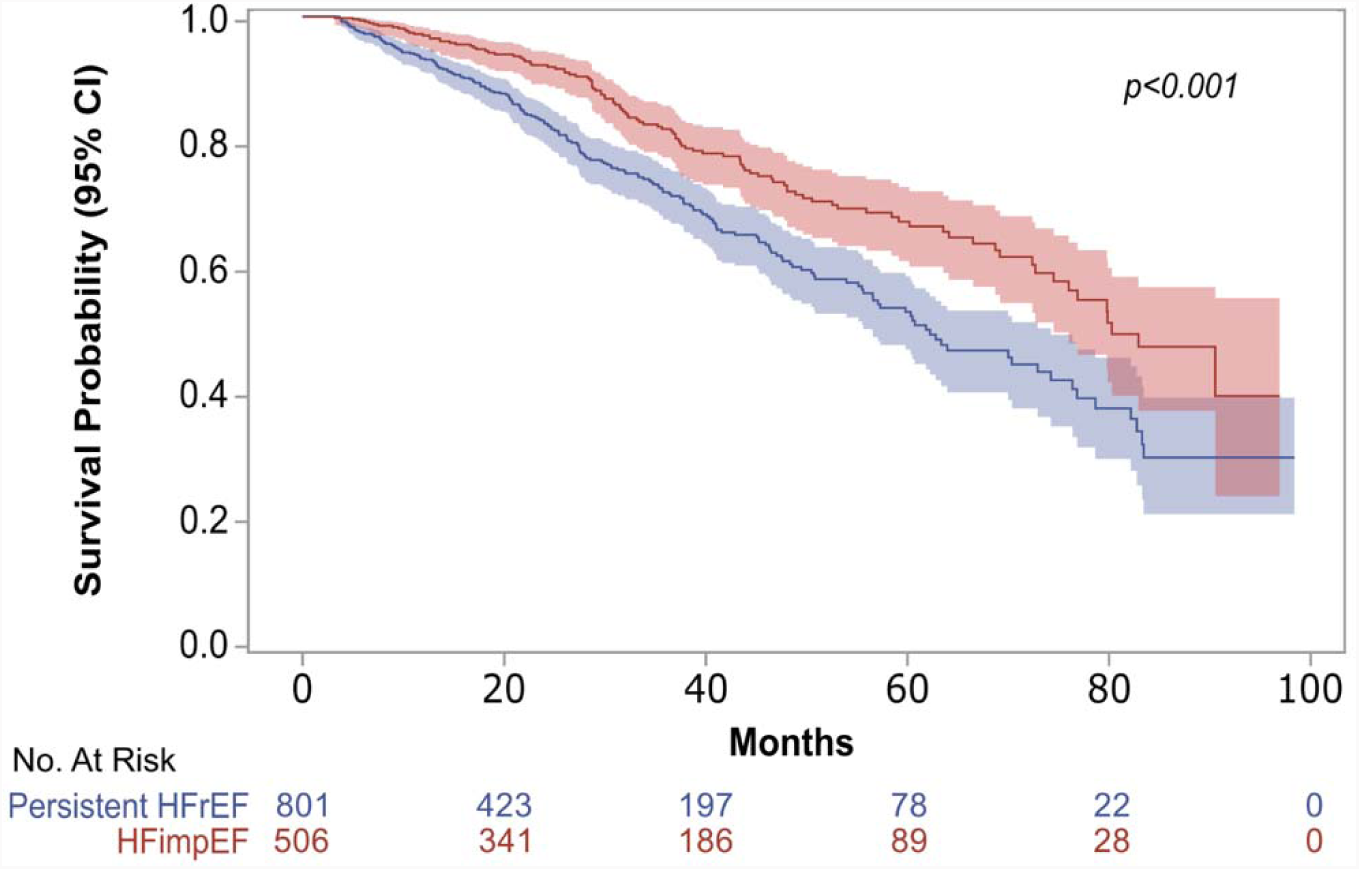
Kaplan-Meier survival curve for all-cause mortality (secondary outcome) by persistent HFrEF versus HFimpEF group.

### Longitudinal trajectories of heart failure

Echocardiogram trajectories are depicted in **Figure 4**. For the HFimpEF group, there was a marked LVEF improvement within the first year, with a trend to decline over time. Conversely, the persistent HFrEF group showed a continuous trend in LVEF decline. Overall, the time-dependent echocardiogram parameters demonstrated that the HFimpEF group had an increasing trend in LVEF, IVSd, posterior wall, and tricuspid annular plane systolic excursion (TAPSE). Meanwhile, a decreasing trend was observed in LVID at end-diastole, LVID at end-systole, and pulmonary artery systolic pressure (PASP). Linear mixed models showed statistically significant (*p*<0.001) changes between groups over time for LVEF, IVSd, LVID at end-diastole, LVID at end-systole, and TAPSE. However, posterior wall and PASP showed minimal longitudinal changes, following the same initial trend (*p*=0.397 and *p*=0.067, respectively). The biomarkers and corrected QT trajectories are shown in **Figure 5**. The HFimpEF group demonstrated significant increased levels of sodium and estimated glomerular filtration rate (*p*<0.001) over time. However, after year four, estimated glomerular filtration rate values seemed to demonstrate a decline. At the same time, BNP and corrected QT values showed slight variations over time, following the same initial trend (*p*=0.083 and *p*=0.129, respectively).

**Figure 4.**
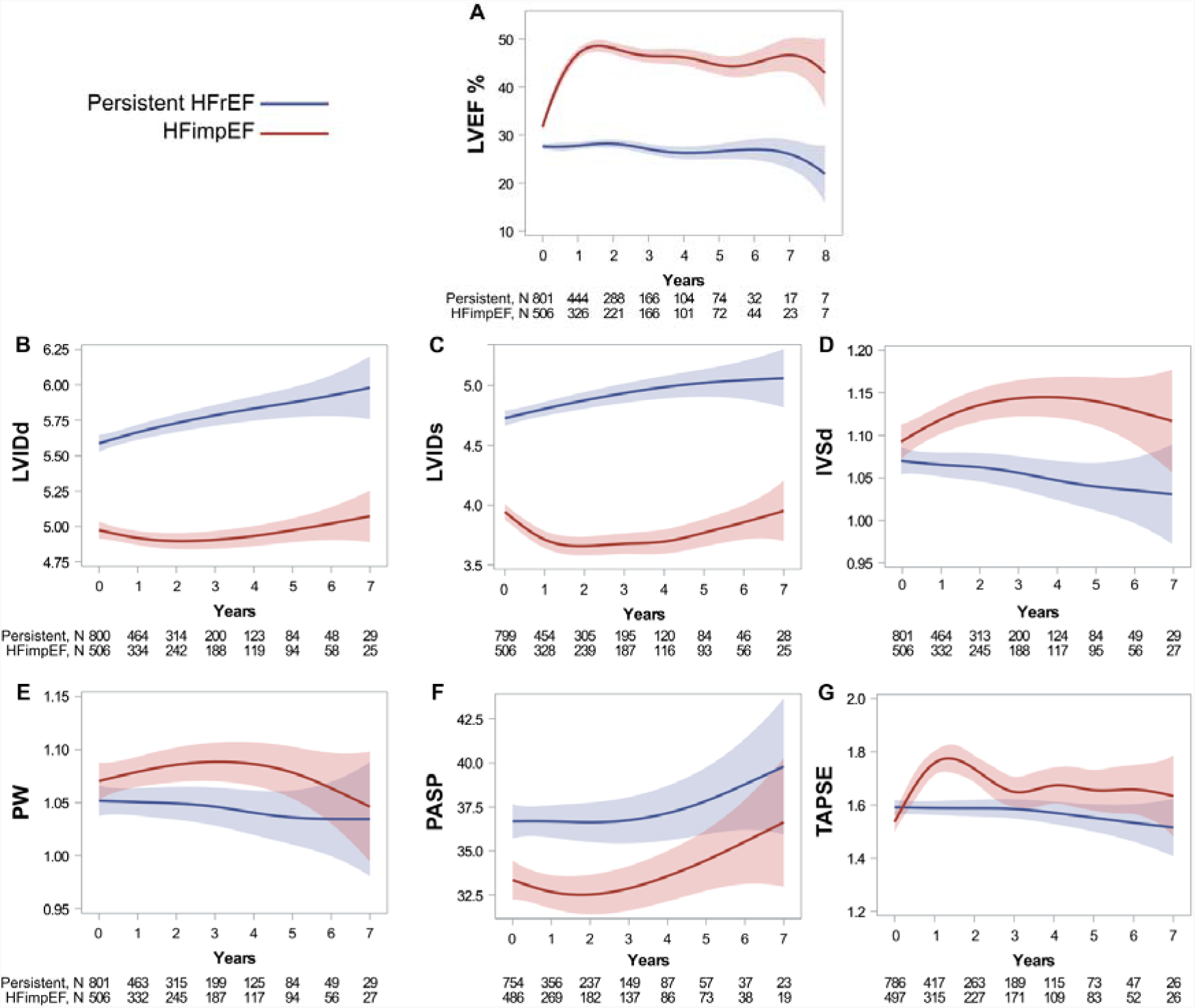
Longitudinal trend of echocardiogram parameters. Data presented as mean (CI) and time as years using penalized B-spline curves. In linear mixed models, LVEF, IVSd, LVIDd, LVIDs, and TAPSE parameters exhibited significant changes over time (*p*<0.001). Only PW and PASP exhibited slight changes, with the same initial trend over time (*p*=0.397 and *p*=0.067 respectively). Abbreviations: IVSd= Interventricular septum thickness at end-diastole; LVEF= left ventricular ejection fraction; LVIDd= left ventricular internal dimension at end-diastole; LVIDs= left ventricular internal dimension at end-systole; TAPSE= tricuspid Annular Plane Systolic Excursion; PASP= pulmonary artery systolic pressure; PW= left ventricular posterior wall.

**Figure 5.**
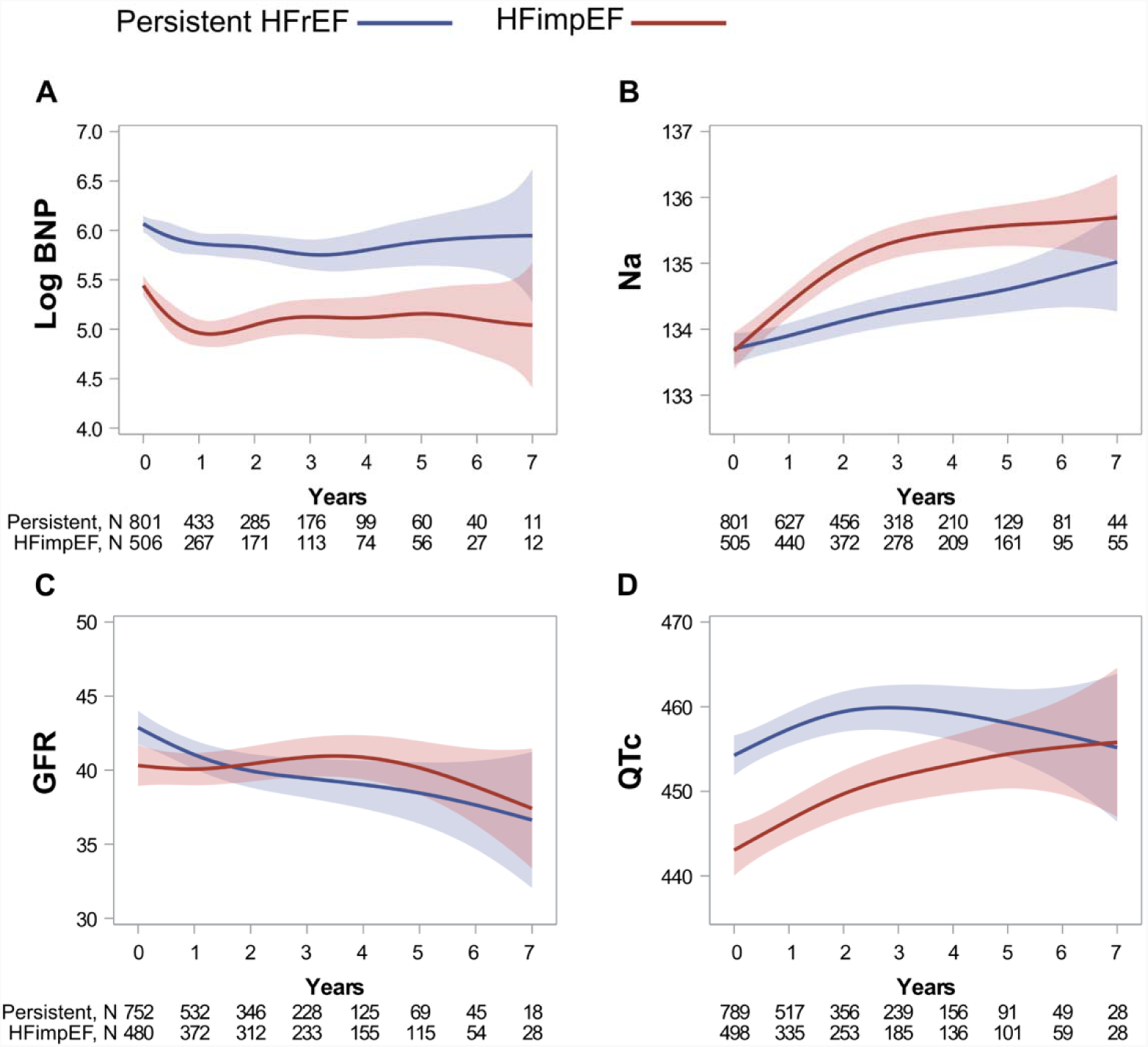
Longitudinal trend of labs and QTc parameters. Data presented as mean (CI) and time as years using penalized B-spline curves. In linear mixed models, eGFR and sodium showed significant changes over time (*p*<.001). Contrarily, BNP and QTC presented the same longitudinal trend without significant changes over time (*p*=0.083 and *p*=0.129 respectively). Abbreviations: BNP= B-type natriuretic peptide; GFR= estimated glomerular filtration rate; Na= sodium; QTc= corrected QT interval for heart rate.

## DISCUSSION

This retrospective HFrEF cohort was conducted to evaluate clinical and longitudinal characteristics associated with HFimpEF. In our HFrEF cohort, it was found that 38% of patients transitioned to HFimpEF and 50% of these patients made this transition within their first follow up year. The HFimpEF Cox regression model identified baseline covariates of persistent or improved HFrEF, which included sex, race, comorbidities, echocardiographic parameters, and BNP. Thus, opening venues for HFrEF improvement risk stratification. Moreover, the HFimpEF group demonstrated distinct echocardiographic and laboratory trajectories and improved survival rates as compared to patients in the persistent HFrEF group.

### Patient baseline characteristics

The baseline characteristics of our HFimpEF cohort consisted of patients who were more likely to be of older age, female, and with increased prevalence of comorbidities such as hypertension and atrial fibrillation. This suggests a non-ischemic profile, which may increase the likelihood of LVEF improvement. In contrast, the persistent HFrEF group demonstrated a higher prevalence of coronary artery disease, higher BNP levels, and less favorable left ventricular geometry and function (left ventricular internal dimensions, posterior wall, IVSd, and LVEF). These characteristics are consistent with previous studies.^4–9^ Lastly, the persistent group exhibited worse echocardiogram and BNP profiles, and they were prescribed RASi and β-blockers more often. This suggests that these treatments were more frequently prescribed to patients with more severe HF.

### Primary HFimpEF endpoint

In the multivariate Cox regression analysis, multiple characteristics were associated with HFimpEF (see **Table 2** and summarized in **Figure 2)**. Of note, in contrast with previous research,^7,8^ associations with HFimpEF were investigated in a time-to-event fashion. These findings could aid in the stratification of patients at risk for developing persistent HFrEF.

Prior research has highlighted that female biological sex is associated with improvement in LVEF.^7,8^ This association was also observed in our multivariate model. Additionally, Black race was related with persistent HFrEF. Previous studies have highlighted that Black patients have an increased susceptibility to structural cardiac remodeling that is more associated with worse systolic function.^16–18^ While in our sensitivity analysis, black race resulted in a week association, these findings support the need for further exploration of the racial disparities underpinning HF among Black patients. Consistent with other HFimpEF studies,^7,8^ patients with atrial fibrillation had an increased likelihood of improving their ejection fraction. It is possible that effective medical or device therapy for controlling atrial fibrillation contributes to this result. On the other hand, patients with coronary artery disease have compromised blood flow to the cardiac muscle with subsequent permanent and irreversible cardiac damage. This type of injury may explain why these patients had a decreased probability of ejection fraction improvement, as has been reported in other studies.^7,8^

An unfavorable left ventricular geometry and function (LVID at end-diastole, IVSd, and LVEF) at the time of diagnosis would be less likely to improve from a mechanistic standpoint. Similarly, elevated levels of BNP can also be a surrogate of unfavorable cardiac function. This may explain the predictive value of these echocardiogram and BNP markers found in our HFimpEF model.

As observed in **Table 1**, β-blockers at <50% of target doses were more frequently prescribed to patients with more severe HF, the persistent HFrEF group. This explains the apparent inverse relationship between β-blockers at <50% of target dose and the HFimpEF outcome. Of note, the retrospective nature of our study does not establish causality. Instead, our findings should be interpreted as associations rather than direct causes and effects.

The RASi and β-blockers at ≥50% target doses were more common in the HFimpEF group (**Table 1**). However, in the multivariate analysis neither were independently associated with HFimpEF (**Table 2**). These findings are consistent with other studies that have reported a lack of association between guideline directed therapy and the definition of HFimpEF.^7,8^ One possible explanation for this could be the enhancement in LVEF by guideline directed therapy in real-world clinical settings is insufficient to meet the established definition criteria for HFimpEF.^3^ Furthermore, guideline directed therapy is frequently underused and under-dosed in health care systems,^19^ as also observed in this study.

Lastly, the persistent HFrEF group was also associated with increased utilization of β-blockers at <50% of target along with lower heart rate and higher prevalence of coronary artery disease. Taken together, this constellation of features delineates a profile of ischemic injury, frequent use of β-blockers, and lower heart rate related to persistently reduced ejection fraction on follow up.

### Secondary mortality endpoint

In the present study, patients in the HFimpEF group demonstrated longer survival rates than their persistent HFrEF counterpart. These results are supported by prior studies demonstrating improved survival rates among patients with HFimpEF.^4–8^ This finding highlights the importance of identifying patients who are at risk of having persistent HFrEF and the need to design and implement interventions to impact outcomes.

### Longitudinal HF trajectories

The longitudinal echocardiographic analysis of the HFimpEF group versus the persistent HFrEF group demonstrated contrasting differences in longitudinal trajectories, most notably, marked LVEF improvement within the first year for the HFimpEF group. Moreover, the HFimpEF group also exhibited increasing trends in LVEF, IVSd, posterior wall, and TAPSE and decreasing trends in LVID at end-diastole, LVID at end-systole and PASP over time. These cardiac geometric and functional changes are indicative of reverse remodeling in the HFimpEF group and remodeling in the HFrEF group.

The values for BNP and corrected QT exhibited small variations over time, following the same initial longitudinal pattern over time between groups. This points towards a persistent electrical remodeling and the persistence of initial BNP levels over time in both groups. Lastly, the HFimpEF group also experienced an increase in sodium and estimated glomerular filtration rate over time, suggesting an improved heart failure leading to renal function enhancement, decrease in neurohormonal activation, and improvement in renal retention of sodium and water. However, despite initial improvements, there was a long-term decline in estimated glomerular filtration rate over time (after four or more years of follow-up).

### Clinical implications

The HFimpEF Cox regression model in the present study has the potential to inform stratification of HFrEF patients who are at risk of experiencing persistent HFrEF at baseline. Early identification of patients at risk for persistent HFrEF creates opportunities to design and implement targeted quality-of-care strategies for guideline-directed therapy, device therapy, timely referrals to advanced HF care, and monitoring strategies to impact HF outcomes.

However, while the model yielded a promising C-statistic of 0.68 given the complexity of the data at hand, validation is required before the model can be implemented for clinical purposes. While prospective randomized studies are ideal for such validation, external and internal validation methodologies can determine reproducibility and generalizability. Additionally, machine learning models could potentially offer more accuracy.

### Future directions in molecular mechanisms

The possible underlying mechanisms for the clinical outcome post insult are likely multifactorial in nature. Inflammatory response activation due to cardiomyocyte loss plays a significant role in the progression of HF towards HFrEF. Cardiomyocyte loss due to apoptosis, necrosis, or pyroptosis is triggered by multiple factors including ischemia, pressure-overload, and mitochondrial dysregulation inducing critical transcriptional factors, inflammatory cytokines and growth factors such as nuclear factor kappa B, tumor necrosis factor, and transforming growth factor-beta, leading to further cardiac fibrosis and cell death.^20–23^ Cardiac fibrosis results in an increase in cardiac stiffness, disruption of coordinated contraction and/or impairment of laminin interaction. Since the underlying comorbidities, life-style modification, as well as the lack of optimal guideline directed therapy can significantly alter the underlying mechanisms, leading to adverse electrical and structural remodeling, therefore, existing comorbidities and medical therapy can have crucial effects on the cardiac outcome trajectory.^24,25^ Furthermore, the intricate balance of ion channels and ionic concentrations has a profound effect on cardiac dysfunction. Intracellular calcium and sodium levels, controlled by sarcoplasmic reticulum Ca^2+^ATPase, Na^+^/ Ca^2+^ exchanger and Na^+^ and Ca^2+^ channels, may contribute to not only to HF phenotype including cardiac arrhythmias, but also mechanistically via transcriptional activation and post-translational modification.^26–28^ To further decipher the myriads of mechanistic underpinnings contributing to HF progression, multiple animal HF models can be utilized.^29^ Due to the similarity to human genome, ease of creating genetic modification, and short breeding cycle, murine models of HF could offer an attractive avenue to study HF, with the ultimate goal of developing novel therapeutic targets. Finally, our study provides insights into how race may feature in HF progression. However, the mechanistic underpinning requires future studies.

### Study limitations

This study has limitations that should be considered when interpreting the results. First, the cohort was derived from a single center, which may limit the generalizability of our findings to other populations and healthcare settings. Second, the observational and retrospective nature of our study precludes causal inferences between the identified predictors and the development of HFimpEF or persistent HFrEF. Our findings should be interpreted as associations rather than direct causes and effects. Additionally, despite adjusting for multiple variables in the multivariate Cox regression analysis, residual confounding due to unmeasured factors cannot be ruled out. Finally, cause-specific mortality analysis is lacking as our institution does not routinely gather cause-specific mortality information.

## CONCLUSIONS

The present study demonstrates that approximately 39% of individuals transitioned to HFimpEF in our HFrEF cohort. Key baseline characteristic predictors in a HFimpEF Cox regression model included sex, comorbidities, echocardiographic measurements, and BNP. If further validated to determine reproducibility, this model could potentially contribute to risk stratification assessment for persistent or improved HFrEF. Early risk stratification enables design and implementation of targeted interventions for quality-of-care improvement impacting HF outcomes. Moreover, the HFimpEF group exhibited better survival rates, and distinct longitudinal echocardiogram and laboratory trajectories when compared with persistent HFrEF.

## Clinical Perspective

### What is new?

- In a retrospective heart failure with reduced ejection fraction (HFrEF) cohort, 39% transitioned to heart failure with improved ejection fraction (HFimpEF), with 50% of these transitions occurring within the first 12 months.
- In a multivariate Cox regression model, sex, atrial fibrillation, coronary artery disease, left ventricular echocardiogram parameters, and natriuretic peptides among others were associated with subsequent development of HFimpEF or persistent HFrEF.

### What Are the Clinical Implications?

- A HFimpEF model could enable risk stratification for persistent HFrEF or HFimpEF.
- If further validated, a HFimpEF model could enable risk stratification for persistent HFrEF or HFimpEF at the time of first indexed ejection fraction ≤40% (index date).
- The risk stratification for persistent HFrEF or HFimpEF has the potential to inform targeted quality improvement strategies, such as guideline-directed medical & device therapies to impact heart failure outcomes.

## Data Availability

All data produced in the study are available upon reasonable request to the authors

## Abbreviations

BNP: Brain Natriuretic Peptide
HF: Heart Failure
HFimpEF: Heart Failure with improved ejection fraction
HFrEF: Heart Failure with reduced ejection fraction
IVSd: Interventricular septum thickness at end-diastole
LVEF: Left ventricular ejection fraction
LVID: Left ventricular internal dimension
PASP: Pulmonary artery systolic pressure
RASi: Renin-Angiotensin-System Inhibitor
TAPSE: Tricuspid annular plane systolic excursion

