## Supplemental Material for "Clinical, Echocardiographic, and Longitudinal Characteristics Associated with Heart Failure with Improved Ejection Fraction"

### SUPPLEMENTARY MATERIAL

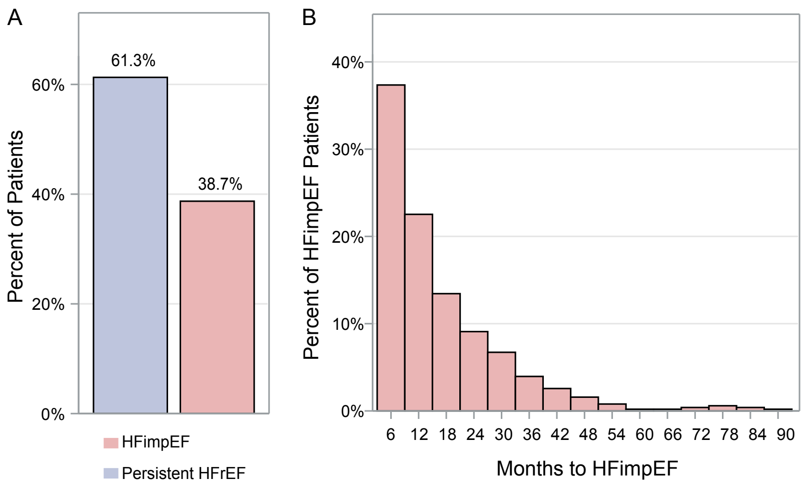


**Figure S1**. (**A**) Proportion of patients from the HFrEF cohort (n=1307) who either transitioned to HFimpEF (n=506, 39%) or persistent HFrEF (n=801, 61%). (**B**) Among the HFimpEF group (n=506), 50% reached the improvement definition within the first 12 months. Histogram representing in the Y-axis the proportion of patients and X-axis representing time to reach the HFimpEF definition.

**Table S1.** Cohort diagnostic performance vs. chart review.

| **Algorithm: ICD codes + LVEF ≤40 + BNP ≥100** | | | | |
|---|---|---|---|---|
| AUC | Sensitivity (CI) | Specificity (CI) | PPV (CI) | NPV (CI) |
| 0.92 | 0.60 (0.48-0.72) | 0.96 (0.91-0.99) | 0.90 (0.80-0.99) | 0.79 (0.72- 0.87) |

Abbreviations: AUC, area under the curve; PPV, positive predictive value; NPV, negative predictive value; CI, confidence interval; ROC, Receiver Operator Characteristic

**Table S2.** Cox regression analyses with Multiple imputation (*m*=50) for the of HFimpEF primary endpoint

| | Multivariate model (no MI) | | Multivariate model with MI[a] | |
|---|---|---|---|---|
| **Variable** | **HR (95% CI)** | ***P* value** | **HR (95% CI)** | ***P* value** |
| Female | 1.31 (1.05-1.63) | **0.017** | 1.35 (1.11-1.64) | **0.002** |
| Race | . | . | . | . |
| Black[b] | 0.74 (0.55-0.99) | **0.043** | 0.80 (0.62-1.04) | 0.096 |
| Other | Reference | . | . | . |
| Atrial fibrillation | 1.48 (1.19-1.84) | **<.001** | 1.51 (1.25-1.82) | **<.001** |
| Coronary artery disease | 0.77 (0.61-0.97) | **0.024** | 0.77 (0.63-0.94) | **0.01** |
| Heart rate (per 20-point increase) | 1.18 (1.07-1.29) | **<.001** | 1.16 (1.07-1.26) | **<.001** |
| β-blocker | . | . | . | . |
| ≥50% | 0.94 (0.74-1.19) | 0.603 | 0.97 (0.76-1.23) | 0.786 |
| <50% | 0.62 (0.47-0.80) | **<.001** | 0.70 (0.54-0.89) | **0.004** |
| None | Reference | . | . | . |
| Log BNP[d] | 0.89 (0.82-0.98) | **0.012** | 0.90 (0.83-0.97) | **0.006** |
| First LVEF ≤40 (per 10-point increase) | 1.20 (1.03-1.40) | **0.018** | 1.21 (1.05-1.38) | **0.006** |
| LVIDd[c] | 0.79 (0.69-0.90) | **<.001** | 0.79 (0.70-0.89) | **<.001** |
| IVSd[c] | 2.17 (1.42-3.30) | **<.001** | 2.08 (1.45-2.98) | **<.001** |

[a]Predictors from the main multivariate analysis that showed a *p* value of ≤.05 were subsequently incorporated into a multivariate analysis with multiple imputation (*m*=50), followed by pooling the derived parameter estimates and associated standard errors.
[b]In the multivariate analysis, race was categorized as black or other.
[c]Hyperlipidemia was omitted in the multivariate model due to possible overlapping effect with coronary artery disease. PW and LVIDs were excluded due to the correlation coefficient >0.5 with IVSd and LVIDd respectively.
[d]The variable has been log-transformed.
**Abbreviations:** BNP= B-type natriuretic peptide; eGFR= estimated glomerular filtration rate; GDMT= guideline-directed medical therapy; IVSd= Interventricular septum thickness at end-diastole; LVEDd= left ventricle end diastolic diameter; LVEF= left ventricular ejection fraction; LVIDd= left ventricular internal dimension at end -diastole; LVIDs= left ventricular internal dimension at end -systole; MAP= mean arterial pressure; MRA= mineralocorticoid receptor antagonist: MI= multiple imputation; NT-proBNP, N-terminal (NT)-pro hormone BNP; PASP= pulmonary artery systolic pressure; PW= left ventricular posterior wall; QTc= QT corrected for heart rate;  TAPSE= tricuspid Annular Plane Systolic Excursion.
